# Cholera deaths during outbreaks in Uvira, eastern Democratic Republic of the Congo, 10-35 months after mass vaccination

**DOI:** 10.1101/2023.05.26.23290528

**Authors:** Patrick Musole Bugeme, Hanmeng Xu, Chloe Hutchins, Juan Dent Hulse, Jaime Mufitini Saidi, Baron Bashige Rumedeka, Moïse Itongwa, Joël Faraja Zigashane Mashauri, Faraja Masembe Lulela, Justin Bengehya, Jean-Claude Kulondwa, Amanda K Debes, Iza Ciglenecki, Esperance Tshiwedi, Faida Kitoga, Tavia Bodisa-Matamu, Taty Nadège, Hugo Kavunga-Membo, Octavie Lunguya, Placide Okitayemba Welo, Jackie Knee, Daniel Mukadi-Bamuleka, Andrew S. Azman, Espoir Bwenge Malembaka

## Abstract

Our understanding of the burden and drivers of cholera mortality is hampered by limited surveillance and confirmation capacity. Leveraging enhanced clinical and laboratory surveillance in the cholera-endemic community of Uvira, eastern Democratic Republic of Congo, we describe cholera deaths across three epidemics between September 2021-September 2023, following mass vaccination.

---

Cholera is an acute diarrheal disease that can rapidly cause severe dehydration and death without prompt and aggressive rehydration[1]. Estimates of the true burden of cholera are highly uncertain because surveillance systems often lack routine identification and testing of suspected cases, and documentation of community cases and deaths is limited. In resource-constrained settings, passive, facility-based surveillance data on mortality are likely to underestimate true mortality; contributing factors include limited patient access to health facilities and incentives for under-reporting[2–5]. Additionally, limited access to confirmatory laboratories for cases and deaths may lead to distorted estimates of the true cholera mortality, both at local and global levels[6]. In the city of Uvira in South Kivu [eastern Democratic Republic of the Congo (DRC)], we implemented an enhanced cholera surveillance system as part of an impact evaluation of preventive mass oral cholera vaccination campaigns conducted in 2020. The killed oral cholera vaccine (kOCV) Euvichol-plus was administered to individuals ≥12 months old living in Uvira in two mass campaigns conducted in July-August and October 2020. Here, we describe cholera deaths across three cholera epidemics in the first three years after vaccination (Figure).

**Figure.**
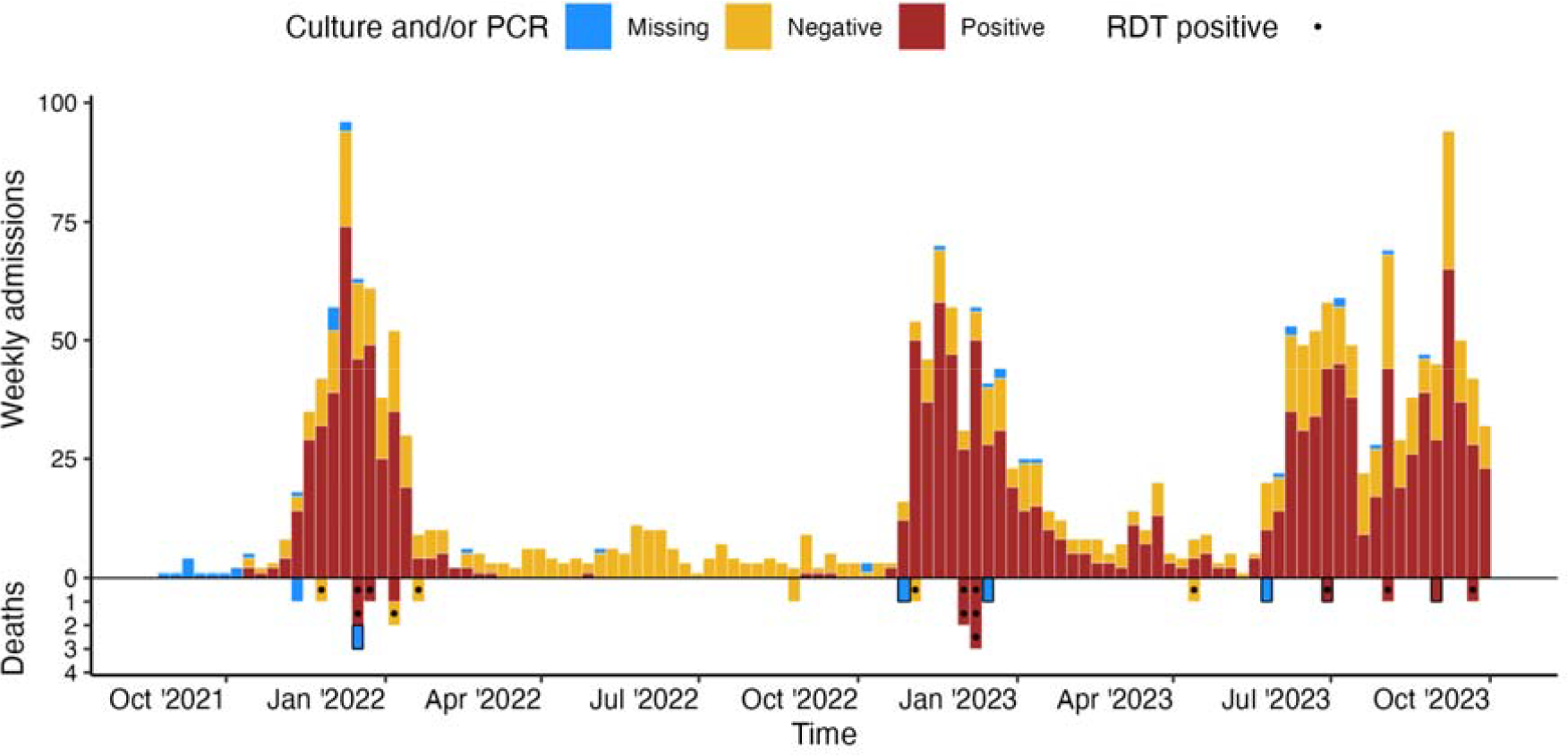
Weekly cholera incidence (top) and death (bottom) by culture or PCR confirmation results in Uvira, 01 September 2021 to 30 September 2023. Before September 10, 2022, culture was performed with a significant lag at an external reference laboratory (INRB Goma) from wet filter papers (stool or rectal swab enrichments suspended in saline), likely leading to reduced sensitivity for detection of *V. cholerae* O1. From September 10, 2022, culture has been performed on-site in Uvira. Rapid diagnostic tests (a mix of O1 and O1/O139 tests) were also used throughout the study. For this reason, we show enriched rapid test positive results as dots to help understand which deaths have more laboratory data supporting *V. cholerae* O1 as the causative agent. Community deaths are highlighted with black boxes.

## Methods

Between September 1, 2021 – September 30, 2023 (10-35 months after the second round of vaccination), suspected cholera cases were recruited at the two cholera treatment facilities in Uvira: the Cholera Treatment Centre at the Uvira General Referral Hospital; and Cholera Treatment Unit at the Kalundu CEPAC health center (both designated as CTC herein). A suspected cholera case was any person ≥12 months old, reporting ≥ 3 acute, watery, and non-bloody diarrheal stools within the 24 hours before hospitalization. Data on sociodemographic characteristics, clinical manifestations, including the level of dehydration based on the Global Task Force on Cholera Control (GTFCC) guidance,[7] vaccination status, and clinical outcome were collected using a structured electronic questionnaire. Written consent was obtained from participants. Rectal swabs and stool samples were collected from consenting patients. Rectal swabs were enriched for 6-18 hours in alkaline peptone water at ambient temperature (∼30°C). Crystal VC Rapid Diagnostic Tests (RDTs; Arkay, India) were used to test raw stool samples and rectal swab enrichments. Samples collected after September 2022 were cultured at the CTC laboratory, whereas those collected before were tested at Rodolphe Mérieux INRB-Goma laboratory in North Kivu. In addition, stool samples from the 2021-2022 outbreak were spotted onto Whatman 903 Protein Saver Cards (Cytiva, UK) and tested by PCR at Johns Hopkins University[8].

Community deaths were not systematically captured by the surveillance system, so the study team learned of suspected community cholera deaths through informal means (Appendix). Visits were organized to collect additional information on the circumstances of death, socio-demographics, clinical manifestations, and vaccination status for each suspected community cholera death. Cholera was the suspected cause of death if the deceased was ≥12 months old and their family members reported that they experienced acute watery diarrhea in the 24 hours preceding the death, with no other cause of death reported. Biological samples were collected on arrival for deaths that occurred during the transit to the CTC.

The characteristics of study participants were compared using the Wilcoxon rank sum and Pearson’s Chi-squared (or Fisher’s exact) tests. We estimated the effectiveness of at least one dose of kOCV against severe cholera (defined by a culture/PCR-confirmed cholera case with severe or treatment plan C dehydration) and suspected cholera death (i.e., any suspected cholera case dying within the CTC with no other cause of death identified) using the screening method, which relies on contrasts of the proportion of cases vaccinated and the vaccine coverage in the population[9]. As vaccine coverage declined over time due to population movement, we estimated a weighted vaccination coverage, based on a log-linear regression of data from three representative vaccine coverage surveys conducted 11, 19 and 29 months after vaccination. We estimated confidence intervals for VE based on a logistic regression model, as previously proposed[9].

## Results

During the 25-month study period, 2209 suspected cholera cases were admitted to the CTCs, of which 1,312 (61.9%) were RDT-positive and 1,460 (67.3%) cases were confirmed by culture or PCR (Table 1). In the same period, 24 suspected cholera deaths were reported, of which 18 (75%) occurred within CTCs, with a facility-based suspected case fatality ratio (CFR) of 0.81% (18/2209).

**Table 1.** Characteristics of patients with cholera who died and survived in the cholera treatment facilities, Uvira, September 2021 to September 2023.

| Characteristic | Overall,<br>N = 2,209 | Suspected<br>cholera deaths, N<br>= 18 | Suspected cholera<br>cases (survivors),<br>N = 2,191 | P value |
| --- | --- | --- | --- | --- |
| Age (years), median (IQR) | 17.0 (7.0,<br>34.0) | 40.0 (21.0,<br>67.8) | 17.0 (7.0, 34.0) | <0.001 |
| Age group (years) |  |  |  | <0.001 |
| < 5 years | 384 (17.4%) | 1 (5.6%) | 383 (17.5%) |  |
| 5-14 years | 608 (27.5%) | 1 (5.6%) | 607 (27.7%) |  |
| 15-59 years | 1,024 (46.4%) | 8 (44.4%) | 1,016 (46.4%) |  |
| ≥ 60 years | 193 (8.7%) | 8 (44.4%) | 185 (8.4%) |  |
| Sex |  |  |  | 0.873 |
| Female | 1,063 (48.1%) | 9 (50.0%) | 1,054 (48.1%) |  |
| Male | 1,146 (51.9%) | 9 (50.0%) | 1,137 (51.9%) |  |
| Health facility |  |  |  | 0.758 |
| CTC (Uvira Referral<br>Hospital) | 1,806 (81.8%) | 16 (88.9%) | 1,790 (81.7%) |  |
| CTU (Kalundu CEPAC<br>health center) | 403 (18.2%) | 2 (11.1%) | 401 (18.3%) |  |
| Level of dehydration on<br>admission* |  |  |  | 0.002 |
| Mild to moderate | 1,060 (48.0%) | 2 (11.1%) | 1,058 (48.3%) |  |
| Severe | 1,149 (52.0%) | 16 (88.9%) | 1,133 (51.7%) |  |
| Time from symptoms onset to<br>hospitalization |  |  |  | 0.253 |
| < 1 day | 1,209 (54.9%) | 9 (50.0%) | 1,200 (54.9%) |  |
| 1 day | 776 (35.2%) | 9 (50.0%) | 767 (35.1%) |  |
| ≥ 2 days | 219 (9.9%) | 0 (0.0%) | 219 (10.0%) |  |
| Missing | 5 | 0 | 5 |  |
| Duration of hospitalization |  |  |  | <0.001 |
| < 1 day | 130 (6.0%) | 7 (38.9%) | 123 (5.7%) |  |
| 1 day | 542 (24.8%) | 6 (33.3%) | 536 (24.7%) |  |
| ≥ 2 days | 1,512 (69.2%) | 5 (27.8%) | 1,507 (69.6%) |  |
| Missing | 25 | 0 | 25 |  |
| Comorbidities** |  |  |  | 0.490 |
| No | 1,449 (91.9%) | 7 (87.5%) | 1,442 (92.0%) |  |
| Yes | 127 (8.1%) | 1 (12.5%) | 126 (8.0%) |  |
| Missing | 633 | 10 | 623 |  |
| Sought care at another health<br>facility |  |  |  | 0.746 |
| No | 1,875 (84.9%) | 15 (83.3%) | 1,860 (84.9%) |  |
| Yes | 334 (15.1%) | 3 (16.7%) | 331 (15.1%) |  |
| Treated at home or pharmacy |  |  |  | 0.557 |
| No | 1,075 (48.7%) | 10 (55.6%) | 1,065 (48.6%) |  |
| Yes | 1,134 (51.3%) | 8 (44.4%) | 1,126 (51.4%) |  |
| Use of antibiotics before admission |  |  |  | 0.232 |
| No | 1,163 (52.6%) | 12 (66.7%) | 1,151 (52.5%) |  |
| Yes | 1,046 (47.4%) | 6 (33.3%) | 1,040 (47.5%) |  |
| APW-enriched RDT |  |  |  | 0.081 |
| Negative | 807 (38.1%) | 3 (17.6%) | 804 (38.2%) |  |
| Positive | 1,312 (61.9%) | 14 (82.4%) | 1,298 (61.8%) |  |
| Missing | 90 | 1 | 89 |  |
| Culture/PCR confirmed cholera |  |  |  | 0.818 |
| Negative | 709 (32.7%) | 6 (35.3%) | 703 (32.7%) |  |
| Positive | 1,460 (67.3%) | 11 (64.7%) | 1,449 (67.3%) |  |
| Missing | 40 | 1 | 39 |  |
| Vaccination status |  |  |  | >0.999*** |
| Not vaccinated | 1,688 (81.0%) | 11 (84.6%) | 1,677 (80.9%) |  |
| At least 1 dose | 397 (19.0%) | 2 (15.4%) | 395 (19.1%) |  |
| Missing | 124 | 5 | 129 |  |

Data are n (proportion) unless otherwise specified. \*: The level of dehydration assessed using the Global Task Force on Cholera Control (GTFCC) guidance.[7] \*\*: Comorbidities refer to self-reported diabetes, hypertension, or HIV infection (Appendix). P values are derived from Pearson's Chi-squared test or Fisher's exact test comparing attributes between survivors and those who died, denoted by \*\*\*. A confirmed cholera case was defined by a positive PCR or culture result. A negative case was confirmed by negative PCR and culture, or by a negative culture only result when PCR was not done.

Fourteen (82.4%) of the 17 health facility deaths in which a sample was collected tested positive for cholera by RDT, and 11 (64.7%) by culture or PCR. The overall culture/PCR-confirmed facility-based CFR was 11 /1449 (0.75%), though this was significantly higher in participants aged ≥ 60 years [4.46% (5/107), p=0.010] compared to younger ones. The suspected cholera CFR in this age-group (≥ 60 years) was 5.45% (11/191) (Table S2).

Comparison between suspected cholera deaths and survivors in health facilities revealed that those dying from cholera tended to be older, with a median (interquartile range, IQR) age: 40.0 (21.0 – 67.8) years] more than double that of survivors [median (IQR): 17.0 (7.0–34.0) years; p < 0.001], with no difference by sex (Table 1). Suspected cases who died were more likely to have been admitted with severe dehydration compared to survivors (88.9% vs 51.7%, p=0.002). Eleven (61.1%) health facility deaths occurred after at least one day of hospitalization. Two of the facility deaths with known vaccination status (n=13), both culture-positive, were reported to have received one dose of oral cholera vaccine during the 2020 vaccination campaign, but we were unable to confirm these with vaccination cards.

The estimated VE was 78.4% (95% CI 74.2–82.4%) against severe culture/PCR-confirmed cholera (dehydration plan C) and 85.5% (95% CI 49.0–97.7%) against death from all-cause diarrhea (suspected cholera death). The method we used to estimate vaccine effectiveness relies heavily on assumptions about the vaccine coverage in the community, and even when assuming a 10% lower vaccination coverage in the population than measured from cross-sectional surveys, the estimated VE against suspected cholera death remained substantial (above 70%) (Figure S1).

The age of community deaths ranged between 4-74 years (median: 38 years), with half (n=3) of them being female. Two community deaths had a stool specimen collected and tested positive for cholera by RDT and culture.

## Discussion

The 0.81% facility-based CFR for suspected cholera cases observed from this passive surveillance system aligns with the WHO target of <1% and was lower than that reported between 2008-2017 in DRC hotspot health zones (1.1%)[10]. While we captured some community deaths, the true cholera mortality burden in Uvira is likely higher than reported here due to lack of robust community-based surveillance and the number of private health facilities that are not integrated into the official surveillance system. The limited existing studies comparing the reported number of facility deaths to those occurring in both facilities and the community suggest this gap is large [2–4,11]. A study from rural Kenya showed that while suspected cholera cases were under-reported by 37%, suspected cholera deaths were under-reported by 200%, implying a 52% underestimate of the community CFR[2–4,11]. In Cameroon, 44% of suspected cholera deaths occurred in the community or during transit to a treatment unit [3]. During the 2010-2011 cholera outbreak in Haiti, community surveys led to almost a 3-fold higher death toll in certain areas compared to official facility-based estimates[4].

A previous report from across DRC suggested that 20.8% of the suspected cholera deaths during the 2008-2017 period were among those under five years old, a stark contrast to the 6.6% in our study [10]. This difference may be a result of several factors including the systematic application of case definitions in Uvira (i.e., improved specificity), differences in health seeking behaviors and/or in the quality of care given to young patients in Uvira.

Almost half of the health facility deaths (8/18) occurred among patients aged at least 60 years, leading to an unacceptably high age group-specific CFR (4.02%). Moreover, 61.1% of deaths happened after ≥ 1 day of hospitalization. This may be because clinical assessment and management of severe dehydration is challenging in the elderly who might have comorbidities requiring slower rehydration pace to avoid fluid overload. Complications related to underlying comorbidities like cardiovascular diseases, diabetes mellitus, anemia, or malnutrition may be overlooked in cholera treatment centers, particularly in the middle of an outbreak, as rehydration is the priority. CTCs in most humanitarian settings, including in our study site, are not sufficiently equipped for an adequate assessment and management of other health conditions cholera patients may present with, including chronic morbidities. Ensuring that CTCs offer comprehensive and person-centered care (as opposed to solely dehydration-centered care) might contribute to the reduction of cholera CFR, particularly among the elderly.

Our findings indicate that the only kOCV currently available in the global stockpile (Euvichol-plus) is highly protective against severe cholera and death. The few available studies reporting kOCV effectiveness against severe cholera point to similar levels of protection, ranging from 73% (23 to 91) 4-24 months post-vaccination in Haiti[12] to 48% (16 to 67) in the fourth year post-vaccination in Bangladesh.[13] Despite the relatively small number of deaths limiting more stratified analyses, this study provides the first, insights into kOCV effectiveness against death and suggests that large-scale deployment of kOCVs in preventive vaccination campaigns may substantially contribute to achieving the Global Task Force on Cholera Control’s goal of reducing cholera mortality by 90% by 2030.[14]

## Conclusion

While the public health community strives to improve the cholera surveillance [15], describing the magnitude and drivers of community and confirmed facility deaths is critical to help correctly target those most at risk and improve patient care and estimates of cholera burden. This study provides a unique insight into confirmed cholera mortality in a resource-constrained and endemic setting. Studies with larger sample sizes, including community surveillance and in different endemicity settings are needed to confirm our findings.

## Data Availability

All data produced in the present work are contained in the manuscript.

