## Appendix for "Cholera deaths during outbreaks in Uvira, eastern Democratic Republic of the Congo, 10-35 months after mass vaccination"

**Web Appendix**

**Table S1. Narrative description of cholera deaths recorded in Uvira, September 2021-September 2023**

| **Cases** | **Brief clinical description** | **Cholera confirmation test** | | |
| --- | --- | --- | --- | --- |
| **Health facility deaths** | | **RDT** | **Culture** | **PCR** |
| Case 1 | A 60–64-year-old male patient was admitted in mid-November 2021 with severe dehydration. He reported watery stools, vomiting, abdominal pain, and fever. He started experiencing the symptoms 4 days before admission and visited a pharmacy where he received metronidazole. The patient died before any samples could be collected for testing. He had not received any cholera vaccine because he was away from home when the vaccinators visited his home. | Missing | Missing | Missing |
| Case 2 | A 45–50-year-old male patient. He was admitted to CTC in the second half of November 2021 with severe dehydration. Before admission, he had experienced watery diarrhea and vomiting one day earlier and was first admitted to the KIGONGO health center where he received an unknown quantity of Ringer’s lactate. Due to his deteriorating condition, he was transferred to CTC where he died on arrival. | Positive | Negative | Missing |
| Case 3 | A 15–19-year-old male patient. He was admitted to CTC in mid-December 2021 with severe dehydration and had experienced watery diarrhea and vomiting for a day. Before admission, he had consulted a prayer home and self-medicated with Metronidazole. He died on the same day of admission.  He did not receive the cholera vaccine during the vaccination in Uvira as he suspected that the vaccine was given against COVID-19 or would contain other microbes. | Positive | Positive | Negative |
| Case 4 | An 85–90-year-old female patient. She was admitted to CTC in mid-December 2021, with severe dehydration following acute watery diarrhea and abdominal pain since the previous day. She did not seek care elsewhere or take medication before admission. She was tested positive by RDT but negative by culture. She died on the second day of hospitalization.  She was not vaccinated against cholera as she believed that the vaccine was given against COVID-19 during the vaccination campaign. | Positive | Negative | Positive |
| Case 5 | A 65–67-year-old female patient. She was admitted to CTC in mid-December 2021. She was severely dehydrated on admission and reported to have experienced tactile fever, watery stools, and abdominal pain since the day of admission. She did not report any self-medication and did not seek care elsewhere. She died on the fourth day of hospitalization.  She was not vaccinated and was residing outside of Uvira at the time of vaccination campaigns. | Positive | Positive | Positive |
| Case 6 | A 65–68-year-old female patient. She was admitted to CTC in early January 2022, with severe dehydration. She reported several episodes of watery diarrhea and abdominal pain several hours before admission. She also reported fever for which she took paracetamol at home. She died within two hours of admission. She reported having been treated for diarrhea in 2005 at a cholera treatment center. She was not vaccinated, saying that the vaccinators did not arrive in her avenue during the vaccination campaign. | Positive | Negative | Negative |
| Case 7 | A 30–34-year-old female patient. She was admitted to the CTC in early January 2022, with severe dehydration following acute watery diarrhea, nausea, vomiting, and abdominal pain. Before admission, she took metronidazole and papaverine) obtained from a pharmacy. She died one day after admission.  She was not vaccinated against cholera because she was absent from the house when the vaccinators went there during the vaccination campaign. She responded herself to the study questionnaire on admission. | Positive | Positive | Positive |
| Case 8 | A 20–24-year-old female patient. She was admitted to the CTC in mid-January 2022, with severe dehydration due to watery diarrhea and vomiting which started on the same day of admission. She did not seek care anywhere else and reported no self-medication. She died one day after CTC admission. She was unvaccinated against cholera because she reported that the vaccinators did not arrive in her avenue during the vaccination in Uvira. | Positive | Negative | Negative |
| Case 9 | An 80–85-year-old male patient. He was admitted to CTC in late August 2022, because of severe dehydration after an acute watery diarrhea that started on the previous day. He also complained of fever and abdominal pain. Prior to his admission, he consulted the BIOMED Polyclinic where he received an infusion of Quinine and other unspecified medications. He received 3500 ml of Ringer’s lactate at CTC but died on the same day of admission. His stool samples tested negative for cholera by both RDT and culture.  He was not vaccinated against cholera as he suspected that the vaccine was given against COVID-19 instead. | Negative | Negative | Missing |
| Case 10 | A female patient under 5 years of age was admitted to the CTC in early November 2022, in a moderate dehydration condition and had experienced acute watery diarrhea for one day. No comorbidity nor self-medication was reported. She received 2000 ml of Ringer's lactate at the CTC but died one day after admission.  She did not receive the cholera vaccine because she was not yet born when the vaccination campaign took place in Uvira. | Positive | Negative | Missing |
| Case 11* | A 15–20–year-old male patient. Before admission, a slight regression of the frequency of watery stools was reported by family members, followed by a worsening of the physical condition by intense asthenia, watery diarrhea, and vomiting. He was self-treated with several medicines (Metronidazole, Ciprofloxacin + Tinidazole and ORS) obtained from a pharmacy before he was admitted to CTC in late November 2022. He was severely dehydrated and died on arrival while resuscitation measures were being put in place (within the first 20 minutes of admission). He received one dose of the cholera vaccine at home during the 2020 vaccination campaign in Uvira, though the vaccination card was no longer available. | Positive | Positive | Missing |
| Case 12 | An 80–85-year-old male patient experienced acute watery diarrhea for one day before he was admitted to the CTU in late November 2022 with severe dehydration. No medication was taken at home before admission. The patient received 12500 ml of Ringer’s lactate, doxycycline and albendazole at the CTU. He died on the 4th day of hospitalization. The patient reported no comorbidities, and he was not vaccinated against cholera because he was living outside the city of Uvira at the time of the vaccination campaign. | Positive | Positive | Missing |
| Case 13 | A 35–40-year-old male patient. He experienced acute watery diarrhea on arrival in early December 2022 and was admitted to the CTC the same day in a profound coma. His clothes were wet and stained with watery stool. He died within minutes of admission when the resuscitation measures were being initiated. The interview was done with close family members who could not provide information on the care-seeking behavior of the patient before admission to the CTC or his vaccination status. | Positive | Positive | Missing |
| Case 14 | An 80–85-year-old female patient. She was admitted to CTC in early December 2022, with severe dehydration. There were no reported comorbidities. She had experienced watery diarrhea, vomiting and abdominal pain one day before admission and self-medicated with an antiemetic (Imodium). She died four days after admission.  She did not receive the cholera vaccine because she suspected that the vaccine was given against COVID-19. | Positive | Positive | Missing |
| Case 15* | A 65–69-year-old female patient was admitted to the CTC in early December 2022 for severe dehydration following a watery diarrhea that started one day earlier. She took metronidazole and other traditional medicines (guava leaves) at home. She died one day after admission, after receiving 7500 ml of Ringer’s lactate at the CTC. She reported to have received one dose of cholera vaccine in July 2020 at a vaccination point in the community in Uvira, but no vaccination card available. | Negative | Negative | Negative |
| Case 16 | A 5–9-year-old male patient was admitted to the CTC in mid-April 2023 with moderate dehydration following acute watery diarrhea and vomiting for one day. Before admission, he self-medicated with Metronidazole and Loperamide (Imodium) at home. He died a day after admission after receiving 3000 ml of Ringer’s lactate. His cholera vaccination status was unknown. | Positive | Negative | Missing |
| Case 17 | A 30–34-year-old male patient who died in early August 2023. He was admitted to the Amani Kwetu health center for watery diarrhea and vomiting one day before death. He received ringer lactate, but treatment was stopped as the family was unable to pay the requested fees. As the level of dehydration of the patient worsened, a transfer to the CTC was requested by the family. The patient arrived dead at the CTC. Rectal swab was collected from the deceased with the consent of his family. His cholera vaccination status was unknown. | Positive | Positive | Missing |
| Case 18 | A 20-24-year-old female patient was admitted in late September 2023 to the CTU for acute watery diarrhea and vomiting with severe dehydration. She took some unidentified medication obtained from a pharmacy before admission. The patient received 13000 ml of Ringer’s lactate and Albendazole at the CTU. She died on the second day of hospitalization. Her mother mentioned that the patient had delayed psychomotor development. Her cholera vaccination status was unknown. | Positive | Positive | Missing |
| **Community deaths** | |  |  |  |
| Case 1 | A 60–64-year-old female patient. The case was reported to the study team by family members who went to the CTC to ask for disinfection materials and assistance for safe funerals as they suspected cholera to be the cause of death. The patient died at her home in mid-December 2021 following a watery diarrhea episode that had begun a few hours before death. She self-medicated with unknown medicines. She was unvaccinated against cholera and no comorbidity was reported. | Missing | Missing | Missing |
| Case 2 | A 65–69-year-old male patient. The case was reported to the study team by his neighbor who was a nurse. He experienced several episodes of watery diarrhea and vomiting in late October 2022, and took some unidentified medication obtained from a pharmacy. She died the same day at home. He was treated again for diarrhea at a cholera treatment center in 2014 and his cholera vaccination status was unknown. | Missing | Missing | Missing |
| Case 3 | A 5–9-year-old male patient. He died in mid-December 2022 after experiencing a watery diarrhea episode and vomiting. This case was reported to the study team by his sister who was admitted to the CTC one day after her brother's death. He had self-medicated then sought care at a local pharmacy. He took Chloramphenicol, Loperamide (Imodium), Metronidazole and ORS before and was admitted to a health facility (the Kilomoni Red Cross health center) where he died on arrival. He was unvaccinated against cholera. | Missing | Missing | Missing |
| Case 4 | A female patient under 5 years of age who died in mid-May 2023 at home after experiencing watery diarrhea and vomiting episodes for a day. This case was reported to the study team by the deceased’s father who was admitted to the CTC two days after that death event. The father tested positive for cholera positive by RDT and culture. The child visited a prayer home and received self-medication with Loperamide (Imodium) and Ringer’s lactate at a local pharmacy before death. Her cholera vaccination status was unknown. | Missing | Missing | Missing |
| Case 5 | A 15–20–year-old female patient who died in late June 2023. The death is likely to have occurred during transit to the CTC. She was reported to have presented with frequent watery diarrhea and vomiting which motivated a visit first to a prayer home, then to an unidentified health center. There she received a perfusion and other medicines of unspecified nature before she was transferred to CTC. She arrived dead. Rectal swab was collected from the deceased and her cholera vaccination status was unknown. | Positive | Positive | Missing |
| Case 6 | A 70–74-year-old male patient who experienced several episodes of watery diarrhea and vomiting in early September 2023. He self-medicated before seeking care at a local pharmacy. He took Metronidazole and ORS, but the persistence of signs and symptoms motivated a transfer to the CTC. On admission, the patient was already dead. A rectal swab was collected from the deceased. Her son reported that she refused the cholera vaccine because of suspicion that the vaccine was given against COVID-19 or could contain other microbes. | Positive | Positive | Missing |

*: Vaccinated with one dose of the oral cholera vaccine. ORS: Oral Rehydration Salts

##### Table S2. Suspected and confirmed cholera cases and case fatality ratio in Uvira

| Population |  | Health facility survivors | Overall CFR (all deaths) | Health facility CFR (health facility deaths) |
| --- | --- | --- | --- | --- |
| **Clinical case definition** | | | | |
| All ages |  | 2,191 | 1.08 (24) | 0.81 (18) |
| 1-4 years |  | 383 | 0.52 (2) | 0.26 (1) |
| 5-59 years |  | 1,617 | 0.68 (11) | 0.55 (9) |
| ≥ 60 years |  | 191 | 5.45 (11) | 4.02 (8) |
| **RDT-confirmed cholera** | | | | |
| All ages |  | 1,495 | 1.12 (17) | 1.06 (16) |
| 1-4 years |  | 243 | 0.41 (1) | 0.41 (1) |
| 5-59 years |  | 1,143 | 0.87 (10) | 0.78 (9) |
| ≥ 60 years |  | 109 | 5.22 (6) | 5.22 (6) |
| **Culture/PCR-confirmed cholera** | | | | |
| All ages |  | 1,449 | 0.89 (13) | 0.75 (11) |
| 1-4 years |  | 236 | 0 | 0 |
| 5-59 years |  | 1,106 | 0.63 (7) | 0.54 (6) |
| ≥ 60 years |  | 107 | 5.31 (6) | 4.46 (5) |

CFR: Case Fatality Ratio, calculated as the percentage of deaths out of all suspected cholera cases (survivors and deaths).

**Vaccine coverage and vaccine effectiveness**

One of the key parameters used for the calculation of the vaccine effectiveness with the screening method is the estimated vaccination coverage in the population [[1]](https://paperpile.com/c/suwMcf/j1Sk). **Figure S1** shows how this parameter can influence the estimated vaccine effectiveness.

**Figure S1. Relationship between population vaccination coverage and vaccine effectiveness against cholera death**

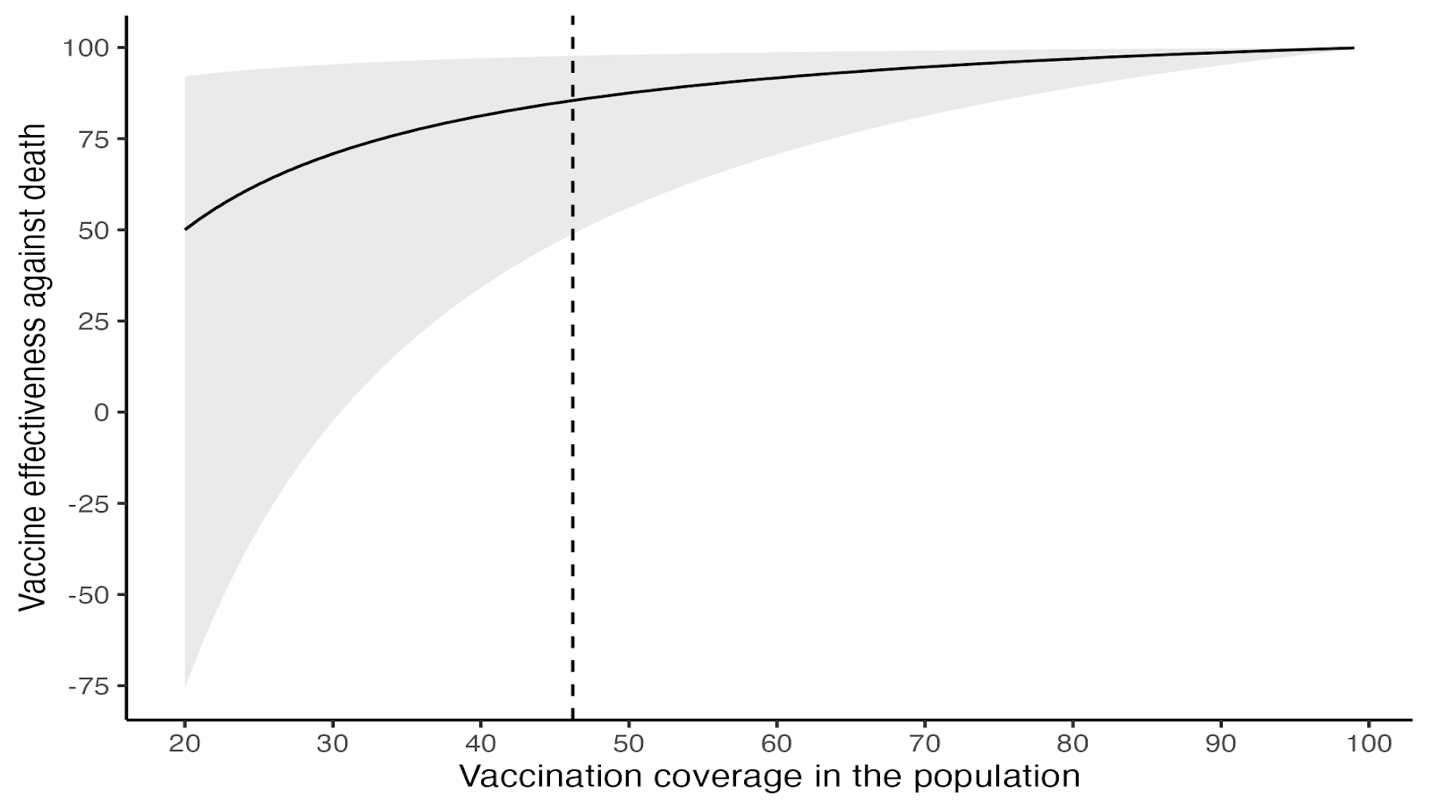

**Figure S1. Relationship between population vaccination coverage and vaccine effectiveness against death.** The dashed vertical line indicates the estimated weighted mean coverage of ≥1 dose of kOCV in the population (46.3%) during the study period (10-35 months after the second round of the 2020 mass vaccination campaigns). The solid line and shading show the estimated vaccine effectiveness and 95% confidence interval, respectively.
